# Urine metabolome of tuberculosis patients receiving intensive phase of treatment show diurnal variations

**DOI:** 10.1101/2021.03.30.21254606

**Authors:** Hossain Md. Faruquee, Heikrujam Nilkanta Meitei, Anupama Pandey, Falak Pahwa, Maria Thokchom, Athokpam Sonia, Shweta Chaudhary, Dinesh Gupta, Huidrom Lokhendro Singh, Sunita Haobam, Reena Haobam, Ranjan Kumar Nanda

**Author notes:** **Corresponding authors:** Ranjan Kumar Nanda (Ph.D.), Translational Health Group, International Centre for Genetic Engineering and Technology, New Delhi-110067, India, E mail and/or Dr. Reena Haobam, Department of Biotechnology, Manipur University, Canchipur, Imphal-795003, Manipur, India, E mail and/or Dr. Sunita Haobam, Department of Pathology, Jawaharlal Nehru Institute of Medical Sciences, Porompat, Imphal-795005, Manipur, India, E mail.

## Abstract

Diurnal variation in biofluid metabolome as observed in a healthy human may alter in perturbed conditions. Biofluids like urine are rich in molecular constituents including metabolites, and infectious disease conditions like tuberculosis (TB) may influence diurnal differences for which limited reports are available in the literature. In this study, we present an optimized gas chromatography coupled to a quadrupole mass spectrometry (GC-MS) method to analyze processed and trimethylsilyl (TMS) derivatized urine metabolites. Urine samples were collected at four time points (0, 6, 12 and 24 hours) of study subjects [n=15; mean age 37 (24-70) in years] including controls [n=7; mean age 29.3 (24-35) years] and culture-confirmed active TB patients [ATB; n=8; mean age 43.7 (25-70) years] receiving treatment in the intensive phase. Global urine metabolite profiling was carried out using the optimized GC-MS method. Higher urine analyte diversity was observed in ATB patients (74) than in controls (36) during the day. Diurnal variations of the parent anti-TB drugs and their breakdown products (pyrazinamide, pyrazinoic acid, 5-hydroxy pyrazinoic acid, isonicotinic acid and alpha amino butyric acid) were observed with maximum abundance at 6 h. Interestingly, urine of ATB subjects at 6 h showed the highest metabolic diversity, whereas it was at 12 h in controls. Many analytes including glycine and alanine amino-acids showed diurnal variation in ATB and controls. These changes could be attributed to the altered host metabolic activities due to disease, treatment-associated decrease in total body bacterial burden and gut microbiota dysbiosis. And the optimized spiked-in internal standard, urine sample volume and GC-MS method could be used for global urine metabolome analysis in healthy and different perturbed conditions.

**“For Table of Contents Only”:** 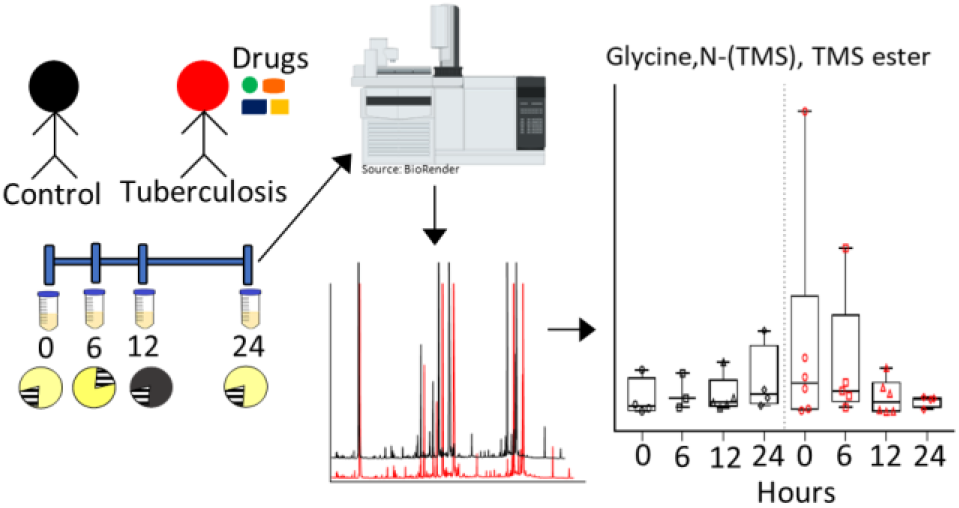

## INTRODUCTION

Circadian clocks control the diurnal rhythm of sleep-wake, feeding-fasting, or rest-activity cycles in mammals.^1,2^ Different biological processes are maintained by the diurnal rhythm which modulates metabolism.^3–5^ Healthy organisms maintain metabolic homeostatic controls at multiple molecular levels.^6^ In perturbed conditions like infectious, metabolic and cardiovascular diseases in human and model organisms demonstrate changes at the tissue-specific transcript, protein and metabolite levels.^7,8^ Similarly, biofluids enriched with metabolic, proteome and lipid components have shown diurnal variations.^9,10^ Reports demonstrated circadian variation in ~15% of the plasma (majorly fatty acids) and saliva (majorly amino acids) metabolites in humans.^11^ Although urine provides a non-invasive collection method from all age groups, limited urine metabolomics studies explaining diurnal variation in healthy or perturbed conditions like infectious disease are available in the literature.^12,13^

Tuberculosis (TB) disease is caused by *Mycobacterium tuberculosis* infection and is the leading cause of mortality, with 1.4 million annual deaths.^14^ TB patients with drug-susceptible Mtb infection receives an intensive two months of treatment with a combination of isoniazid (INH), rifampicin (RIF), ethambutol (ETB) and pyrazinamide (PZA) followed by four months of continuation phase including RIF and INH.^15^ TB patients experience loss of appetite, loss of muscle mass and body weight. Metabolite profiles of sputum, serum and urine of TB patients have shown deregulated molecular pathways.^16^ Earlier report has shown the presence of rhythmicity in biofluid metabolites of healthy and different disease conditions.^17–19^ However, diurnal variation in urine metabolites, if any, of TB patients receiving medication, is yet to be systematically studied using metabolomics, which might bring additional details for a better understanding of perturbed pathophysiology in TB.

Commonly, nuclear magnetic resonance (NMR) and gas or liquid chromatography and mass spectrometry (G/LC-MS) are employed for global biofluid metabolite profiling.^20,21^ Mass spectrometry methods yield wider coverage and GC coupled to quadrupole MS instruments provides a cost-effective tool for metabolite profiling. And a standardized GC method might be useful to researchers having access to GC-MS to employ and monitor diurnal variation to comprehend changes in different disease conditions. Detailed molecular analysis in perturbed conditions with focus on diurnal variations will bring new knowledge in understanding disease processes.^22^

In this study, we report an optimized spiked-in internal standard, urine sample volume and GC-MS method to identify diurnal variation in urine metabolome of controls and TB patients. This method could be used to monitor anti-TB drugs and their breakdown products in TB patients receiving therapy. The diurnal variation of urine metabolites in TB patients, as observed, could be attributed to altered feeding pattern, host metabolism, treatment-associated decrease in total body bacterial burden, disease and treatment-induced gut microbiota dysbiosis. This method could be employed by researchers involved in infectious disease biology like TB and chrono-biology to identify disease-specific markers which may have a translational potential in adjunct therapeutic development.

## EXPERIMENTAL SECTION

### Study subject recruitment

This study was performed in accordance with the protocol approved from the Institutional Ethics Committee of Jawaharlal Nehru Institute of Medical Sciences (JNIMS), Imphal (Ref No. Ac/04/IEC/JNIMS/2017); Manipur University, Imphal (Ref No. Ac/IHEC/MU/003/2017) and International Centre for Genetic Engineering and Biotechnology (ICGEB), New Delhi (Ref No. ICGEB/IEC/2018/06). After receiving signed consent form, subjects presenting with at least 2 weeks of cough, weight loss, fever symptoms at outpatient department of JNIMS, Imphal were recruited. Sputum samples, day 1 random and day 2 early morning, were collected and subjected to sputum microscopy, a cartridge based nucleic acid amplification test and culture tests. Subjects with positive test results, for the above tests, were included in the study as active tuberculosis patients (ATB) and the rest were excluded. Procedures followed for subject recruitment, classification, sample collection and processing are presented in Figure 1. Asymptomatic healthy subjects, not receiving any medication for at least 3 weeks, were recruited as controls. Adult subjects (>18 years) with negative human chorionic gonadotropin (hCG) urine test results were included in this study. ATB patients received an anti-TB drug daily regimen of INH (300 mg), RIF (600 mg), PZA (1500 mg) and ETB (900 mg) following the standard recommendations of the Revised National TB Control Programme (RNTCP) of India. After at least two weeks of receiving TB drug regimen in the intensive phase, mid-stream urine samples (~ 40 mL) from these TB patients were collected at different (0 h, 6 h, 12 h and 24 h corresponding to 06:00-08:00, 12:00-14:00, 18:00-20:00 and 06:00-08:00 respectively) post daily drug dose. Urine samples from healthy control subjects were collected following similar method for comparison. Urine samples, after collection were stored at 4 °C and within 6 hours of collection, were shifted to −80 °C at clinical sites till further analysis.

**Figure 1.**
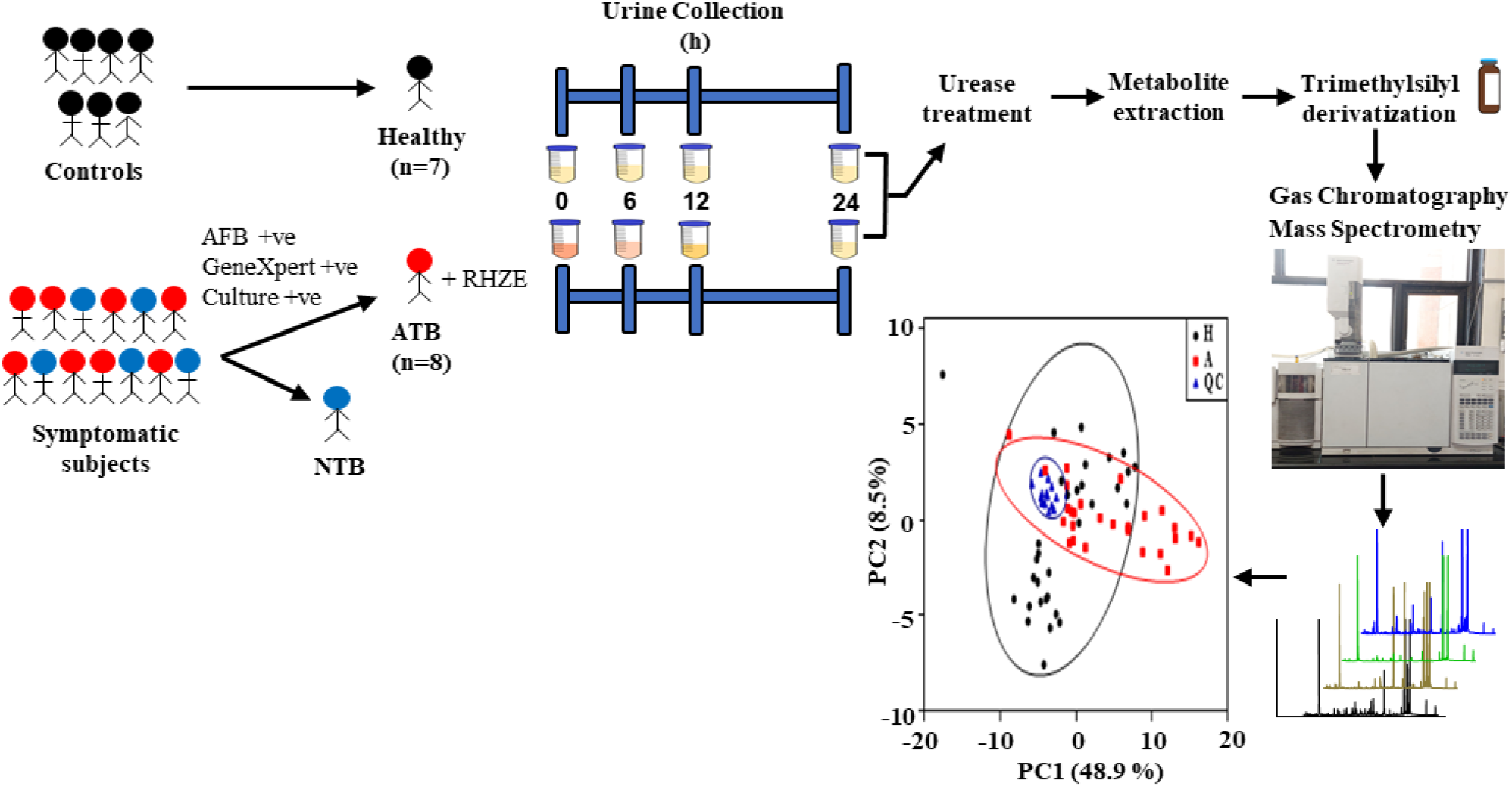
Schematic representation of the steps followed for study subject recruitment and classification for urine sample collection, processing and metabolite profiling using Gas Chromatography-Mass Spectrometry (GC-MS). ATB: Active tuberculosis patients; NTB: Non-tuberculosis; RHZE: rifampicin (R), isoniazid (H), pyrazinamide (Z), ethambutol (E); PC: Principal component; H: Healthy control; A: ATB; QC: Quality control; h: hours.

### Pre-processing and standardization of urine samples for metabolite extraction and derivatization

Stored samples were thawed on ice and given a short spin at 4 °C for 2 min. A quality control (QC) sample was prepared by pooling equal volumes of urine samples from all ATB subjects (n=8) and their time points for standardization experiments. QC samples (100 µL) with internal standard (ribitol) were treated with urease (0.5 U/µL) following earlier reported methods.^23^ Different amounts of ribitol (2.5, 5, 10, 20 and 40 µg; equivalent to 25, 50, 100, 200 and 400 ng injection volume) were added in QC and incubated in a dry bath at 37 °C for 1 h. Metabolite extraction was carried out by adding methanol (800 µL), followed by centrifugation at 10,000 × *g* at 4 °C for 10 min. The supernatant (~ 900 µL) was transferred to a fresh 1.5 ml microcentrifuge tube and dried using SpeedVac (Labconco, USA) at 40 °C for 3-4 h. Toluene (100 µL, dried over Na_2_SO_4_) was added to the dried pellet and vacuum dried until complete dryness. To the dried samples, N, O-Bis (trimethylsilyl) trifluoroacetamide (BSTFA; 100 µL) with 1% chlorotrimethylsilane (TMCS) reagent was added and incubated at 60 °C for 1 h in a thermomixer (Eppendorf, Germany). The TMS derivatized urine metabolites were centrifuged at 10,000 × *g* for 5 min and 80 µL of the supernatant was transferred to a glass vial insert (200 µL) in a GC vial (2 mL capacity) for GC-MS analysis. Similarly, different volumes of urine QC samples (50, 100, 150, 200, and 250 µL) and standardized amount of ribitol (10 µg) were processed for further analysis. Optimized urine volume from both study groups (control: n=7, ATB: n=8) at all time points were taken for pre-processing, metabolite extraction, derivatization and GC-MS data acquisition. Similar pre-processing and derivatization methods were followed for commercial standards of anti-TB drugs and their breakdown products; isoniazid (INH), acetyl-isoniazid (AcINH), isonicotinic acid (INA), isonicotinuric acid (INTA), pyrazinamide (PZA), pyrazinoic acid (POA), 5-hydroxy pyrazinoic acid (5OH-POA), ethambutol (ETB), and alpha-amino butyric acid (AABA) (10 µL, 10 mg/mL) and amino acids (10 µL, 10 mg/mL; Sigma, USA) prior to GC-MS analysis.

### Gas chromatography-mass spectrometry data acquisition

GC-MS analysis was performed using a 7890 Gas Chromatography coupled to a quadrupole mass spectrometry (Agilent Technologies, Santa Clara, CA). The TMS derivatized standards or test urine metabolites (1 µL) were injected to an RTX-5 column (5% diphenyl, 95% dimethylpolysiloxane; 30 m × 0.25 mm × 0.25 μm; Restek, USA) in a splitless mode. Helium was used as a carrier gas at a constant flow rate of 1 mL/min. Electron ionization (EI) mode was fixed at −70 eV to scan ions of 33 to 600 m/z range. A solvent delay of 6.15 min was used. The ion-source temperature and quadrupole temperature were fixed at 230 °C and 150 °C respectively. Sample introduction to data acquisition parameters were controlled through ChemStation software (Agilent Technologies, USA). The front inlet temperature was fixed at 250 °C; and six sets of GC method parameters were used to select an optimized GC-MS method to acquire maximum coverage; GC1: 50 °C for 1 min, ramp of 10 °C/min to 150 °C, hold for 3 min, ramp of 7 °C/ min to 300 °C, hold for 3 min (run time 38.43 min); GC2: 50 °C for 1 min, ramp of 10 °C/min to 150 °C, ramp of 5 °C/ min to 300 °C, hold for 3 min (run time 44 min); GC3: 120 °C for 0 min, ramp of 4 °C/min to 300 °C, hold for 2 min (run time 47 min); GC4: 120 °C for 0 min, ramp of 8 °C/min to 300 °C, hold for 2.5 min (run time 25 min); GC5: 50 °C for 1 min, ramp of 5 °C/min to 150 °C, hold for 2 min, ramp of 7 °C/ min to 300 °C, hold for 3 min (run time 47.43 min); GC6: 50 °C for 1 min, ramp 7 °C/min to 300 °C, hold for 3 min (run time 39.71 min). The MS scan speed was used in three different modes: normal (2.59/sec), fast (12.95/sec), and very fast (15.68/sec). Out of the different GC-MS methods, GC1 method with normal scan speed was used for the rest of the test sample data acquisition. A derivatized QC sample was run using the optimized method six times to calculate the relative standard deviation (RSD). All test samples were randomized using online tool (www.randomizer.org) and were run in multiple batches (10 samples per day with at least two QC before starting and ending the batch GC-MS runs). Data acquisition of derivatized samples was completed within 24 hours of processing.

### Data Processing

All GC-MS raw data files were converted to .cdf files in MSD ChemStation for transferring and processing them with LECO@ChromaTOF software (4.50.8.0, CA, USA). For peak picking, peak width was set at 0.5 s and the signal to noise ratio (S/N) threshold was 75. NIST (National Institute of Standards and Technology, USA) library (version 11.0, mainlib and replib including 2,76,248 total spectra) for putative analyte identification, and library identity search with a similarity match of >600 at a mass threshold of 100, was used. Acquired data files of QC samples and test urine samples from control and ATB subjects were aligned using “Statistical Compare” feature of ChromaTOF. The .csv file generated after feature alignment with peak areas and S/N information was selected for data matrix preparation. Manual curation was followed to check the identity of the analytes and those with less similarity (<600) to the library spectra, duplicates, silanes and analytes with S/N <75 were removed from the data matrix. To maximize the identification of the anti-TB drug molecules and their breakdown products, low stringent criteria (10% of analytes present in the four time points of all subjects) was used. The analytes that were absent in more than 50% of samples of at least one study group were later excluded from the analysis.

### Chemometric analysis

The data matrix was manually normalized to the total peak area and taken for further chemometric analysis using MetaboAnalyst 5.0.^24^ Internal standard normalized data (80 variables, after removing 5 variables related to parent or breakdown products of anti-TB drugs; 3 groups: QC; n=16; four time points data of control: n=7 and ATB: n=8) was selected for Principal Component Analysis (PCA) to find the clustering pattern. In subsequent analyses, separate matrix considering variables present in at least 50% per time point in control (36 variables × 7 × 4 time points) and ATB (74 variables × 8 × 4 time points) subjects were selected to identify study group specific important analytes that may explain altered diurnal variations, if any. Missing values of variables were filled with half of the minimum positive values within the group. Generalized log transformation (glog_2_) with auto-scaling was selected and a Partial Least-Squares Discriminant Analysis (PLS-DA) model was built using the control and ATB data matrix separately. Analytes with a Variable Importance in Projection (VIP) score of >1 were selected as important. To identify the common and unique analytes in urine samples of control and ATB subjects, Venny 2.1.0 tool was used.

### Statistical analysis

Two-tailed t-test was employed to calculate differences in age and body mass index (BMI) of the study subjects. Individual analytes and their normalized peak areas were used to prepare box and whisker plots (mean values, minimum to maximum with all points) using GraphPad Prism (7.00, GraphPad Software, San Diego, CA). PCA and PLS-DA score plots generated from MetaboAnalyst 5.0 were redrawn using GraphPad Prism Software. All data points were plotted as mean ± standard error of the mean.

## RESULTS

### Study Groups

The study subjects from control and ATB [n=15, 67 % male, mean age 37 (24-70) years] groups showed a significant difference (p=0.0072) in their BMI (Table S1). A lower BMI of ATB patients was observed as compared to controls. Also, the majority of ATB patients were smokers and alcohol users.

### Optimization of GC-MS parameters and sample volume for urine metabolite analysis

Earlier reported methods were used in extracting urine analytes and subsequent derivatization.^23^ An attempt was made to optimize methods to acquire data using commonly available quadrupole GC-MS in place of a multidimensional GC-MS. We observed that 100 ng of ribitol, as a spike-in standard, provides enough signal in urine metabolite analysis (Figure 2A, Figure S1). From the different GC-MS methods used for data acquisition (GC1-GC6), GC1 (50 °C for 1 min, ramp at 10 °C/min to 150 °C, hold for 3 min, ramp at 7 °C/ min to 300 °C and hold for 3 min) yielded maximum urine analyte details (Figure 2B, 2C, Figure S2). Employing the optimized GC-MS method parameters, amongst metabolites extracted from different volumes of QC samples, 200 µL urine sample volume generated optimum information and was selected for data acquisition of all test samples afterwards (Figure 2D). Data matrix was prepared using LECO@ChromaTOF software and the total analytes in the metadata reduced from 605 to 85 after following the set criteria. The selected analytes (~85) covered amino acids, fatty acids, organic acids, mono-saccharides, hormones and other molecular classes (Table S2, Figure S3).

**Figure 2.**
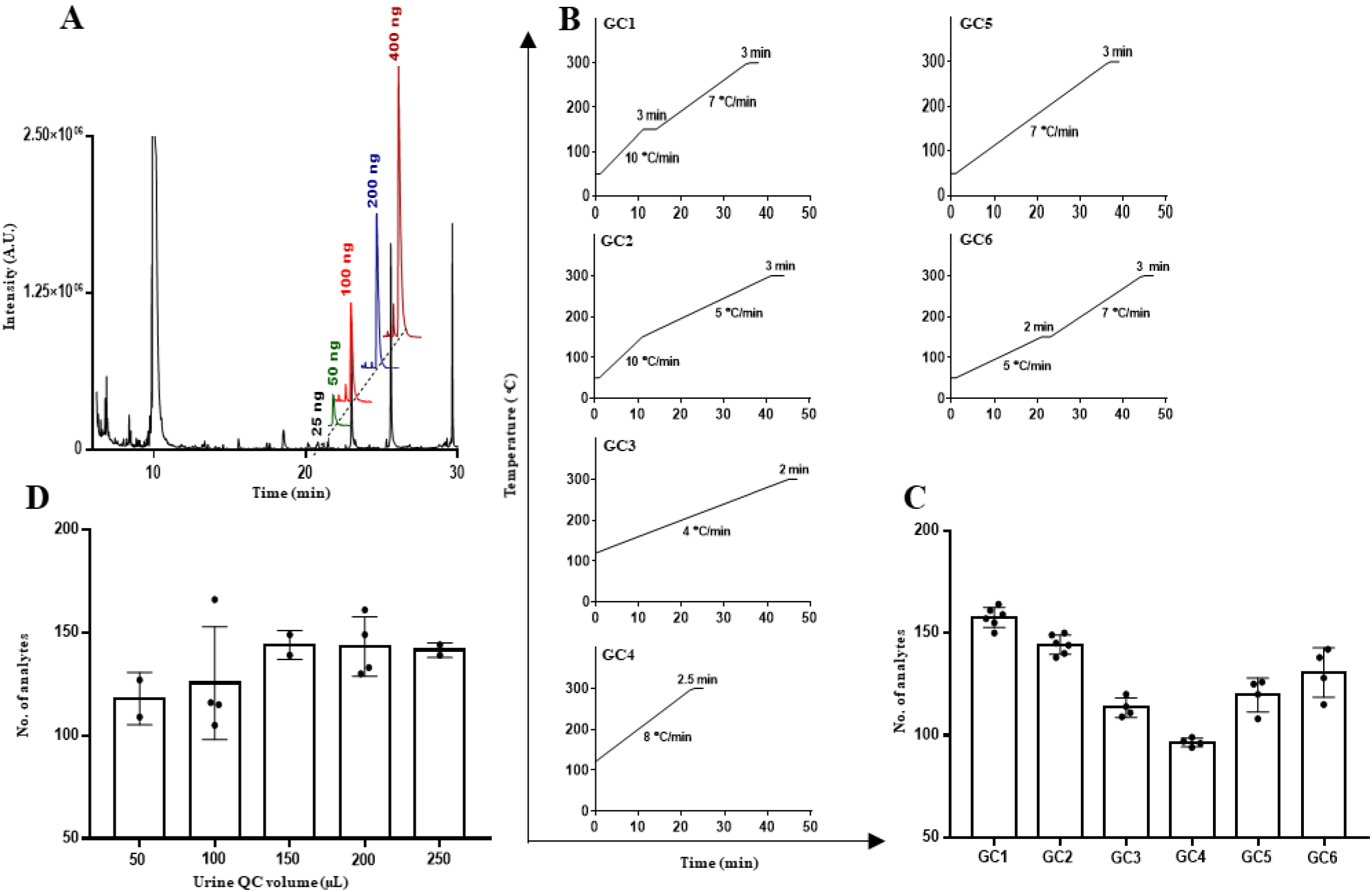
Optimization of internal standard amount, gas chromatography (GC) parameters and sample volume for global urine metabolite profiling. **A**. Different ribitol amount (25, 50, 100, 200 and 400 ng) was profiled using GC-MS. **B**. Different sets (GC1-GC6) of GC parameters followed for profiling urine analytes. **C**. Number of urine analytes identified using different sets of GC parameters. **D**. Number of urine analytes from different QC volumes as identified using the optimized GC-MS parameter (GC1). A.U.: arbitrary units, min: minutes.

### Tuberculosis drug distribution in urine of ATB patients receiving treatment

In ATB patients, parent anti-TB drugs and their breakdown products: PZA (POA and 5 OH-POA), INH (INA) and ETB (AABA) were identified in their urine samples (Figure 3A–3D, Figure S4). Each of these analytes showed diurnal variation, following a time-dependent trend, and the majority showed the highest abundance at 6 h post receiving anti-TB drug dose.

**Figure 3.**
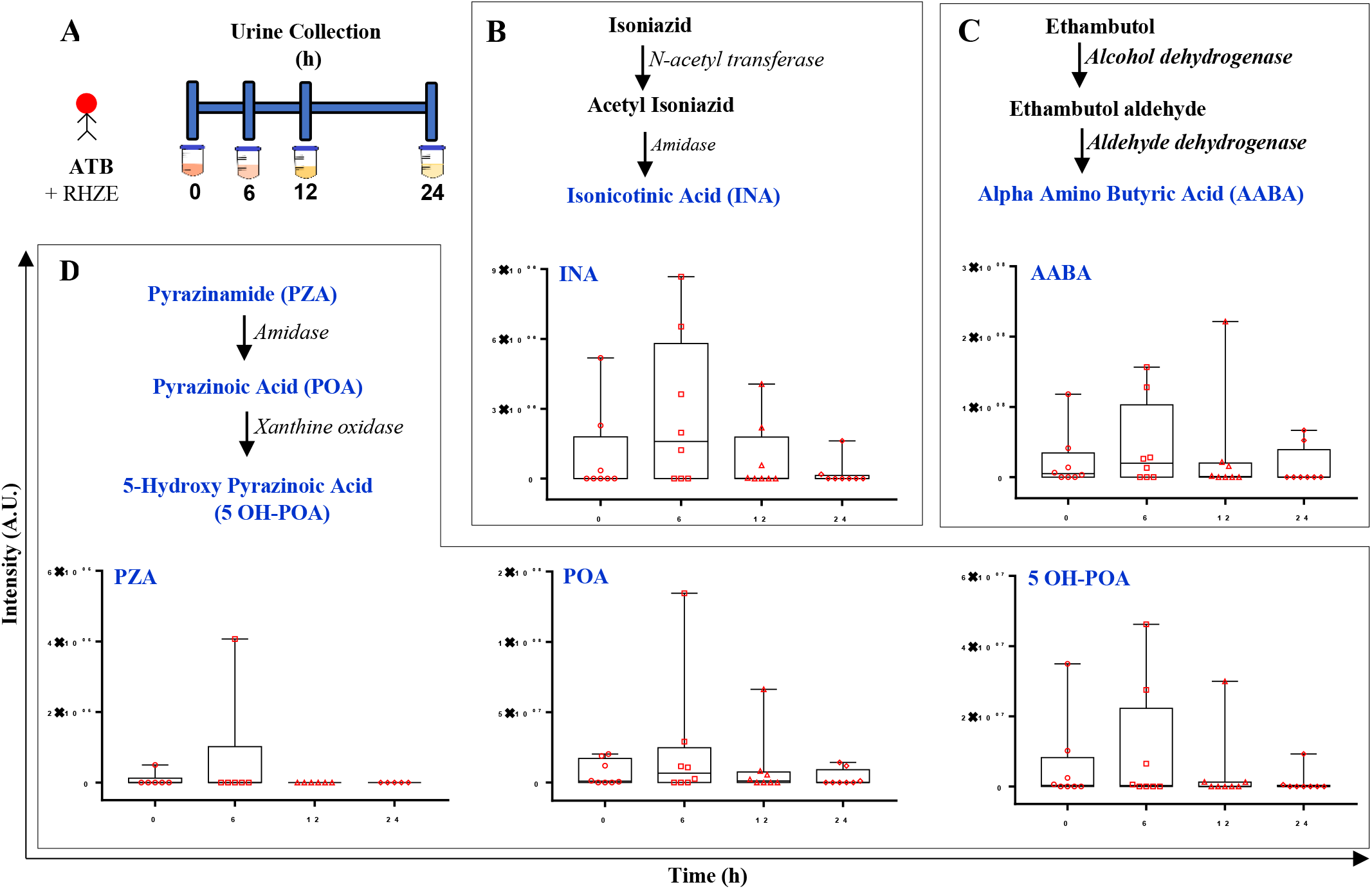
The abundance of anti-tuberculosis (TB) drugs and their breakdown products varies at different time point post-dose urine samples of ATB patients. **A**. Urine sample collection at four time points (0, 6, 12 and 24 h) from TB patients receiving treatment. **B, C, D**. Highest abundance of anti-TB drugs and their breakdown products were observed at 6 h post receiving the dose in majority of ATB patients. RHZE: rifampicin (R), isoniazid (H), pyrazinamide (Z), ethambutol (E). A.U.: arbitrary units, h: hours.

### Global urine analytes in controls and TB patients

The urine analytes identified in the control study group showed that 19 out of 36 analytes were present in all four time points, with highest number at 12 h (Figure 4A). Similarly, 25 out of 74 identified analytes in ATB subjects in all time points wherein 6 h samples had the highest number of analytes (Figure 4B). A shift in molecular diversity in ATB subjects to an earlier time of the day with respect to control was observed. Out of these two sets of analytes in ATB and controls, we observed 13 analytes were common in both study groups and the rest were unique to each of them (Figure 4C). The individual abundance of these 13 common analytes at different time points of the day showed a time dependent trend between control and ATB subject groups (Figure 4D). The majority of these common analytes showed the highest abundance at 6 h in the healthy subject group except hexopyranose, which was found to be the lowest (Figure 4D). Similarly, ATB subjects had a majority of these common analytes (6) showing the highest abundance at 24 h, four at 12 h and three at 0 h (Figure 4D). All the unique molecules identified in control and ATB subjects showed a diurnal variation and peaking at different time points of the day (Figure S5).

**Figure 4.**
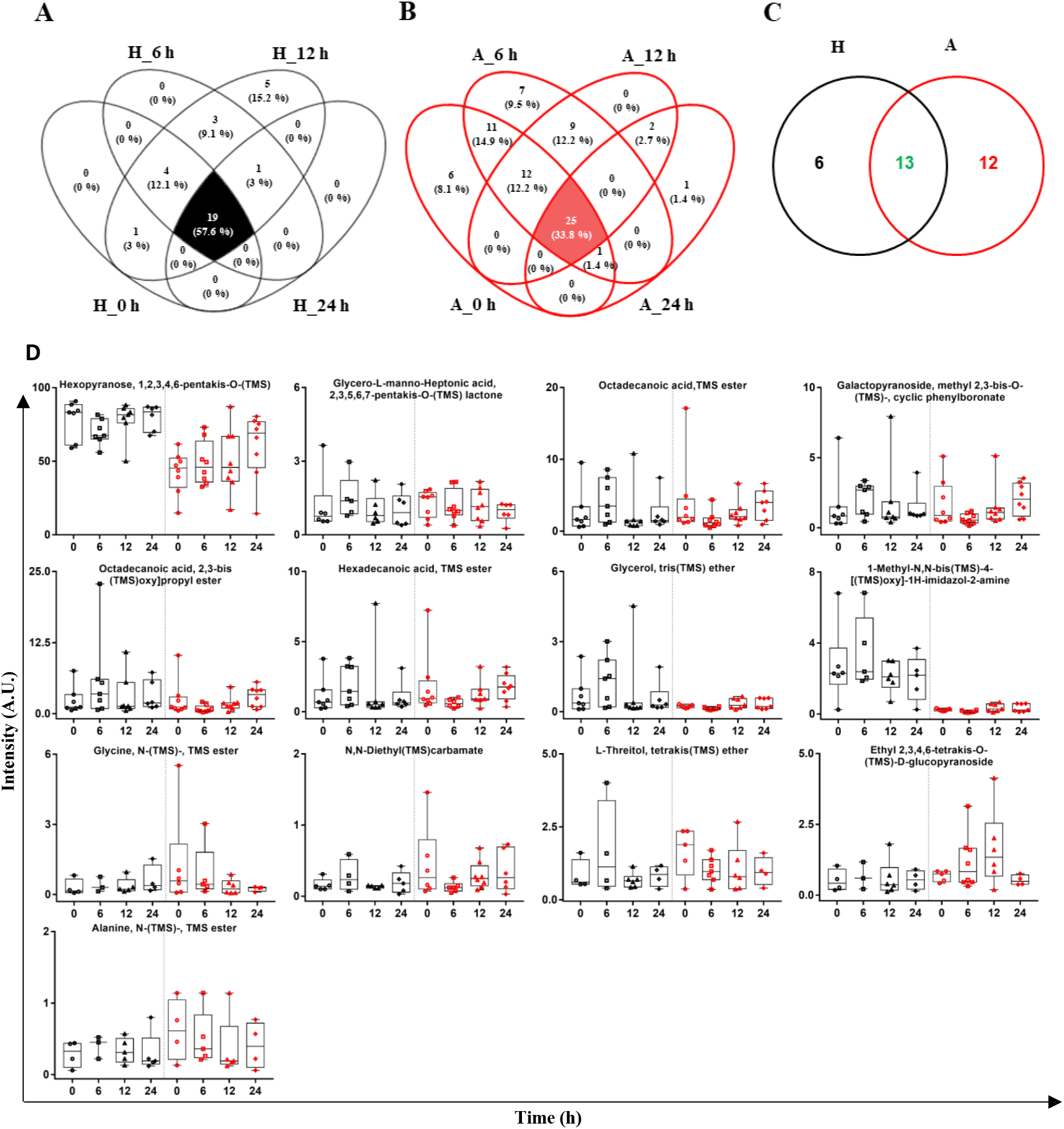
Urine metabolites identified in control and tuberculosis (TB) subjects showed diurnal variation and higher diversity in ATB patients. Venn diagram showed common analytes identified within and between the control and ATB subjects at different time points. **A**. Urine analytes identified at different times of the day in healthy control group. **B**. Urine analytes identified at different times of the day in ATB patients. **C**. Common and unique urine analytes identified in all time points of the control and ATB subjects. **D**. Individual common analytes (13) showed significant diurnal variation with majority in control (black) and ATB (red) having the highest abundance at 6 h and 12 h respectively. A.U.: arbitrary units; h: hours.

### Diurnal variation in urine metabolome of TB patients and controls

Identified urine metabolites (n=80) in two study groups (control: 7 and ATB: 8) at four time points and all QC samples (n=16) clustered closely (Figure 5A). Samples of both the study groups showed partial overlap in the PCA plot. PLS-DA model, including metabolites as variables, time points as groups and urine samples as observations in ATB patients (n=8, 32×74), showed overlapping clusters between time points (Figure 5B). A similar observation was also observed in PLS-DA model including urine metabolite metadata (27×36) of control subjects (n=7) (Figure 5C). A set of 9 metabolites was found to qualify a VIP score >1 each in both ATB and control study groups (Figure 5D, 5E) and two molecules (1,2,3-Propanetricarboxylic acid, 2-[(TMS)oxy]-, tris (TMS) ester and Galactopyranoside, methyl 2,3-bis-O-(TMS)-, cyclic phenylboronate) were common. Urine Propanetricarboxylic acid was found to follow a pattern between control and ATB subjects with the lowest at 12 h and at 6 h respectively (Figure 5F). In contrast, 2,3-methyl Galactopyranoside showed an inverse trend and reached maximum at 6 h in control but at 24 h in ATB subjects (Figure 5G). At 6 h in control subjects, most of the identified important analytes showed the highest abundance (Figure 5H). And five out of the rest were found to be minimum at 6 h in ATB groups (Figure 5I). Abundance of hexopyranose was found to increase during the day, with the highest at 24 h in ATB groups (Figure 5I). The majority of these molecules showed a time-dependent change in their abundance at different times (0, 6, 12, and 24 h) of the day and the rest remain unchanged. Urine Alanine and Glycine showed a change in their diurnal abundance in control and ATB subjects. The identity of these analytes was confirmed by running commercial standards (Figure S6).

**Figure 5.**
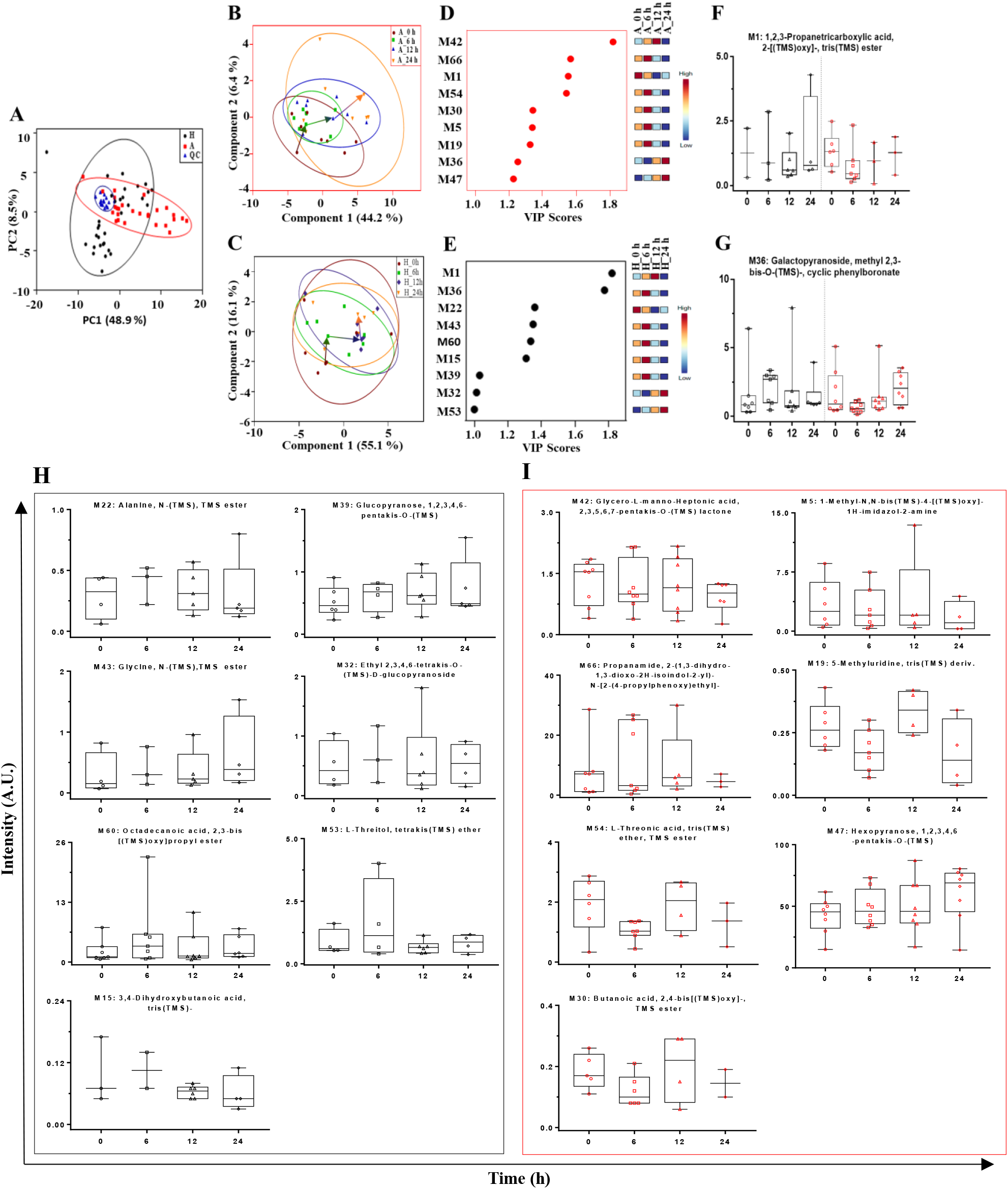
Identification of important urine analytes showing diurnal variations in control and tuberculosis (TB) subjects. **A**. Principal Component Analysis (PCA) of quality control (QC), control (H: Healthy) and active TB (A: ATB) samples showed a close clustering of all QC samples. Partial Least Square Discriminant (PLS-DA) model built using urine analytes of all time points of control (**B**), and ATB (**C**) subjects. Diurnal variation of a representative subject’s follow-up urine samples from H and A is manually connected by arrows (**B, C**). Analytes with VIP score >1 in control (**D**) and ATB subjects (**E**). Individual abundance of two common (**F, G**) and seven unique (**H, I**) molecules from control (black) and ATB (red) subjects each at different times of the day. A.U.: arbitrary units; h: hours.

## DISCUSSION

Many biological processes follow circadian rhythms, driven by internal and external clues that synchronize and coordinate organ physiology and behaviour.^25–27^ In infectious disease conditions like TB, patients report a decreased appetite and higher muscle mass loss due to increased catabolism. All these pathophysiological changes impact the host metabolism and may reflect in host biofluid composition. Global metabolites in biofluids of TB patients have been analyzed and deregulated sets of molecules were identified and reported in the literature.^23,28,29^ Earlier reports from our team and others have identified anti-TB drugs and their breakdown products in the urine of TB patients receiving treatment.^23,30^ However, a standardized method using GC coupled to a quadrupole MS might be beneficial to the researchers due to its wide accessibility which could be adopted to identify important analytes in the urine of patients with different physiological and metabolic disorders.^31,32^

Ribitol was used as an internal standard and 100 ng of it was found to be sufficient in global urine metabolite analysis. Earlier optimized urine sample processing methods to remove urea by urease treatment and TMS derivatization steps prior to GC-MS data acquisition were adopted without any changes.^23^ After following and comparing different sets of GC parameters and MS scan modes, GC1 method (50 °C for 1 min, ramp at 10 °C/min to 150 °C, hold for 3 min, ramp at 7 °C/ min to 300 °C and hold for 3 min) and 2.59/sec scan speed separated urine analytes well in a 38.43 min run. On average, 160 analytes were identified in the QC samples representing all ATB samples. Although, urine sample collection involves a non-invasive method and is easy for patients of all age groups, higher volumes of starting material may lead to a crowding effect, hampering resolution during molecular profiling in GC-MS analysis. Hence, in our optimization experiments, we observed that 200 µL of urine sample yields the highest molecular details (~160 analytes). A QC sample was run consecutively six times using the optimized GC-MS method and showed an RSD of <3.52% suggesting the method is reproducible. Identified analytes belonged to different molecular classes like amino acids, fatty acids, monosaccharides, organic acids and others which could be of the host, pathogen and/or gut microbiota origin. During the day, urine osmolarity changes based on the hydration levels hence total area normalization was carried out in our analysis. Creatinine level is commonly used as a normalization factor in urine metabolite analysis, but in perturbed conditions like TB, due to muscle mass loss, creatinine level is reported to be higher, so was not considered in this study. ^26,33,34^

Earlier, we monitored urine metabolites in TB patients just after receiving the first dose of treatment^23^ but conditioning the host system needs at least a few weeks before the enzymes responsible for TB drug metabolism gets stabilized. So, we collected urine samples from subjects completing at least 15 days post-therapy. Urine samples were collected at four time points (0, 6, 12 and 24 h) from recruited ATB subjects receiving anti-TB drug dose (for at least 15 days) in a day post-dose where 0 h corresponds to 6:00-8:00. We could identify a few parent anti-TB drugs and their breakdown products in the urine samples of ATB subjects using the optimized GC-MS method. The majority of these analytes showed diurnal kinetics with the highest abundance at 6 hours post receiving drug dose and comport previous findings.^23,28^ Many of the anti-TB drugs and their breakdown products in urine could be identified using higher starting sample volume (~1.5 mL) or adopting a GC×GC-TOF-MS method as reported earlier.^23^ However, our aim was to capture the diurnal changes in urine analyte composition and their abundance in TB patients.

Interestingly, even after removing anti-TB drugs and their breakdown products, the total number of urine analytes was always higher in ATB subjects (ATB=74, controls=36). In control subjects, ~53% of the analytes were present at all time points, whereas ~34% were common in ATB patients. We observed changes in total urine analyte composition and abundance in TB patients and controls at different time points of the day, which demonstrates altered pathophysiology and higher biofluid metabolic diversity in TB patients receiving treatment.

It is important to note that reports suggest early morning urine samples are rich in protein and nitrogenous substances and afternoon specimens are better suited for electrolyte analysis.^33^ However, our observation in TB patients indicated the highest metabolic analytes at 6 h (12:00-14:00) and at 12 h (18:00-20:00) in controls. Energy expenditure is usually higher during the day in healthy subjects. Diurnal variations of tryptophan breakdown to nicotinamide were observed in women, which increased significantly with cold stress.^36^ Our results suggest a probable shift in the energy expenditure pattern and maxima in TB patients on which limited reports are available. So, with altered pathophysiology in TB patients, a shift in diurnal variation to earlier times of the day with the highest urine analytes was observed.

In all time points, a total of 13 analytes was found to be common in both TB and controls in this study. It is interesting to note that most of these common analytes show the highest abundance at mid-day (6 h) in TB subjects and at evening (12 h) in controls. Urine glycine level was highest at 6 h in controls, whereas it was highest at 0 h in ATB patients. Urine glycine is reported to get altered significantly in sleep-deprived conditions and in subjects suffering from depression.^1,37^ In TB patients undergoing therapy, depression has been reported which might have an impact on treatment adherence.^38,39^ Six unique analytes (Glucopyranose, Mannofuranuronic acid, 3,4-Dihydroxybutanoic acid, Propanoic acid, Phenyl acrylate and Glycine) were identified in controls and twelve (2H-Indazole, Butenedioic acid, Talose, Methylmandelic acid, 5-Methyluridine, Fructofuranose, N-Acetyl-D-glucosamine, Mannose, 1,2-dithiane, Acetic acid, Benzeneacetic acid and Fucopyranose) in TB patients. Understanding the biological functions of these analytes, after validating their identity, would further explain the increased number of unique analytes in the urine of TB patients.

To understand the processing, derivatization, GC-MS method and instrument-associated variations, at least two QC samples were run with the study samples in each batch. All QC samples clustered closely together in PCA analysis, assuring that the method-associated changes were minimal. Separate PLS-DA analyses of all time point samples of controls and ATB groups, showed that early and late time point samples cluster with a high degree of overlap. Two important molecules were common in both study groups with minimum abundance of 1,2,3-Propanetricarboxylic acid at 12 h in controls and at 6 h in ATB. Galactopyranoside abundance was highest at 6 h in controls whereas at 24 h in ATB. The rest of the important unique analytes in control showed the highest abundance at 6 h. Similarly, 4 out of 7 analytes in ATB subjects (Glycero-L-manno-Heptonic acid, 5-Methyluridine, L-Threonic acid and Butanoic acid) showed the highest levels at 12 h. Alanine in control showed the highest diurnal level at 6 h while increased glycine levels were observed during the day. The presence of pathogen causes a significant alteration in the pathophysiology of TB patients and is reflected as increased urine metabolic diversity. Both amino acids are components of Mycobacterial cell wall and used directly from host for synthesizing its biomass. Previous report highlighted a deregulation of tyrosine and phenylalanine amino acid metabolism in TB patients.^16^

Inter-individual differences in the urine samples of ATB and healthy controls were observed. This change could be attributed to food habit, age, environmental exposure, treatment-induced decrease in total host body Mycobacterial load, altered gut microbiota associated with disease and/or treatment conditions out of several other factors.^40,41^ The control subjects were younger than the ATB subjects. However, as the study subjects were from the same geographical place, they follow similar food habits and environmental exposure and therefore, may have contributed to minimum variation. Similar but controlled experiments, including TB patients and controls receiving equal total food energy and exposure conditions (ambient temperature, humidity) might be useful to generate interesting information on diurnal variation at molecular levels. Correlating the total body Mycobacterial burden and urine metabolome has recently yielded interesting conclusions.^34^ Diet, physical activity and hormones play major roles in diurnal rhythmicity and are responsible for fluctuations. ^42,43^ So, maintaining uniformity in time of urine sample collection and hormonal status are critical in comparative metabolomics and epidemiological studies.

Although our study confirmed earlier reported findings and added new details on the diurnal differences in urine analytes of TB patients, it has several limitations. Primarily, the study sample size is small and we and others^44^ have used control subjects that are not age-matched, which could be treated as an important limitation of the study. Taking these findings to a population level needs additional studies, with case and controls from similar age groups. The effect of different phases of TB treatment, like continuation phase and its influence on urine metabolite composition and diurnal variation in non-responders to treatment were not considered in this study. Similarly, TB patients infected with drug-resistant and diverse resistant cases might contribute to a certain degree of variation. However, our results elucidating that TB patient urine samples are more diverse than healthy subjects and show altered diurnal pattern, are still significant. Many of the urine metabolomics studies employ random or early morning sample collection methods, but as observed in this study, the timing of sample collection could influence the conclusion. These shortcomings could be taken care with more extensive and focused studies in different countries simultaneously considering appropriate sample size and patients receiving similar duration of treatment. A focused programme based on multi-institutional and multi-centric studies, including subjects from a different continent, will expand our findings and yield important outcomes for further refinement.

## CONCLUSIONS

We have optimized internal standard, urine sample volume and a GC-MS method for a global urine metabolome analysis with high reproducibility. Employing the optimized method, we observed the highest abundance of identified anti-TB drugs and their breakdown products at 6 hours post-dose during the intensive phase of treatment. Interestingly, TB patients’ urine samples were found to show higher molecular diversity at different times of the day than controls. The highest number of urine analytes were observed earlier in TB patients (mid-day, 6 h) than in controls (evening, 12 h). Urine metabolomics yield interesting information depicting an altered pathophysiology and a deregulated diurnal variation in TB patients. Detailed understanding of the diurnal differences at population level will yield interesting avenues to monitor and manipulate important host molecules that might be useful for developing adjunct therapeutic options.

## Data Availability

All mass spectrometry data files, used in this report, will be available to users by sending an email to the corresponding author.

## ASSOCIATED CONTENT

### Supporting Information

Figure S1, Identification of Ribitol (internal standard, 100 ng) and spiked-in quality control urine sample (QC); Figure S2, Total ion chromatograms (TIC) obtained from different Gas Chromatography and Mass Spectrometry (GC-MS) methods used for global urine metabolite analysis; Figure S3, TIC showing the peak distribution of different QC volumes used for metabolite extraction, derivatization and GC-MS data acquisition; Figure S4, TIC and retention time (RT) of the derivatized commercial standards of parent anti-tuberculosis drugs and their breakdown products; Figure S5, Unique urine analytes identified in controls and active tuberculosis patients (ATB) receiving treatment showed diurnal variation in their abundance Figure S6, TIC and RT of the derivatized commercial standard amino acids and spiked-in QC.

Table S1, Epidemiological details of study subjects; Table S2, Tentatively identified human urine analytes in healthy control and active tuberculosis patients receiving treatment.

## AUTHOR INFORMATION

### Author Contributions

RH, RKN, DG, HLS, SH conceptualized the project, acquired funding, shared resources and administrated the project. Under the supervision of HLS, SH and RH, subject recruitment, collection of epidemiological details and samples were collected by HNM, AP, MT, AS. HMF, FP completed sample processing, methodology standardization, validation, gas chromatography-mass spectrometry data acquisition. HMF, FP, SC, RKN analyzed the data and wrote the original draft. The manuscript was finally reviewed and edited by HNM, AP, MT, AS, SC, DG, HLS, SH, RH.

### Notes

All authors declare no conflict of interest.

## ACKNOWLEDGMENTS

We would like to acknowledge the contribution of all the hospital staffs at clinical sites involved in tuberculosis patient management and recruitment. Funding: Funding for this work was provided by the Department of Biotechnology (MDR-TB/2017/39), Government of India, as a research grant to RKN, DG, RH, SH, HLS. The core support from International Centre for Genetic Engineering and Biotechnology, New Delhi to RKN is highly acknowledged. FH has an Arturo Falaschi Fellowship support from ICGEB, Trieste, Italy. FP is supported by the fellowship from Department of Biotechnology, Government of India. SC is a Shyama Prasad Mukherjee fellow and is supported by Council of Scientific and Industrial Research, Government of India. Dr. Shyam K. Masakapalli from Indian Institute of Technology, Mandi for going through the manuscript and for providing his valuable comments for improving the manuscript.

## Notes

### Competing Interest Statement

The authors have declared no competing interest.

### Clinical Trial

This is a clinical research project.

### Author Declarations

This study was performed in accordance with the protocol approved from the Institutional Ethics Committee of Jawaharlal Nehru Institute of Medical Sciences (JNIMS), Imphal (Ref No. Ac/04/IEC/JNIMS/2017); Manipur University, Imphal (Ref No. Ac/IHEC/MU/003/2017) and International Centre for Genetic Engineering and Biotechnology (ICGEB), New Delhi (Ref No. ICGEB/IEC/2018/06).

