## Supporting information for "Urine metabolome of tuberculosis patients receiving intensive phase of treatment show diurnal variations"

### AUTHOR INFORMATION

### CORRESPONDING AUTHORS

*
*
*

**Table of contents** **Page**

| Table of contents | Page |
|---|---|
| **Figure S1.** Identification of Ribitol (internal standard, 100 ng) and spiked-in quality control (QC) urine sample. | **S2** |
| **Figure S2.** Total ion chromatograms (TIC) obtained from different Gas Chromatography and Mass Spectrometry (GC-MS) methods used for global urine metabolite analysis. | **S3** |
| **Figure S3.** TIC showing the peak distribution of different QC urine sample volumes used for metabolite extraction, derivatization and GC-MS data acquisition. | **S4** |
| **Figure S4.** TIC and retention time of the derivatized commercial standards of parent anti-tuberculosis drugs and their breakdown products. | **S5** |
| **Figure S5.** Unique urine analytes identified in controls and active tuberculosis (ATB) patients receiving treatment. | **S6** |
| **Figure S6.** TIC and retention time of the derivatized commercial standard amino acids and spiked-in QC urine samples. | **S7** |
| **Table S1.** Epidemiological details of study subjects. | **S8** |
| **Table S2.** Tentatively identified human urine analytes in healthy controls and ATB subjects receiving treatment. | **S9-S12** |

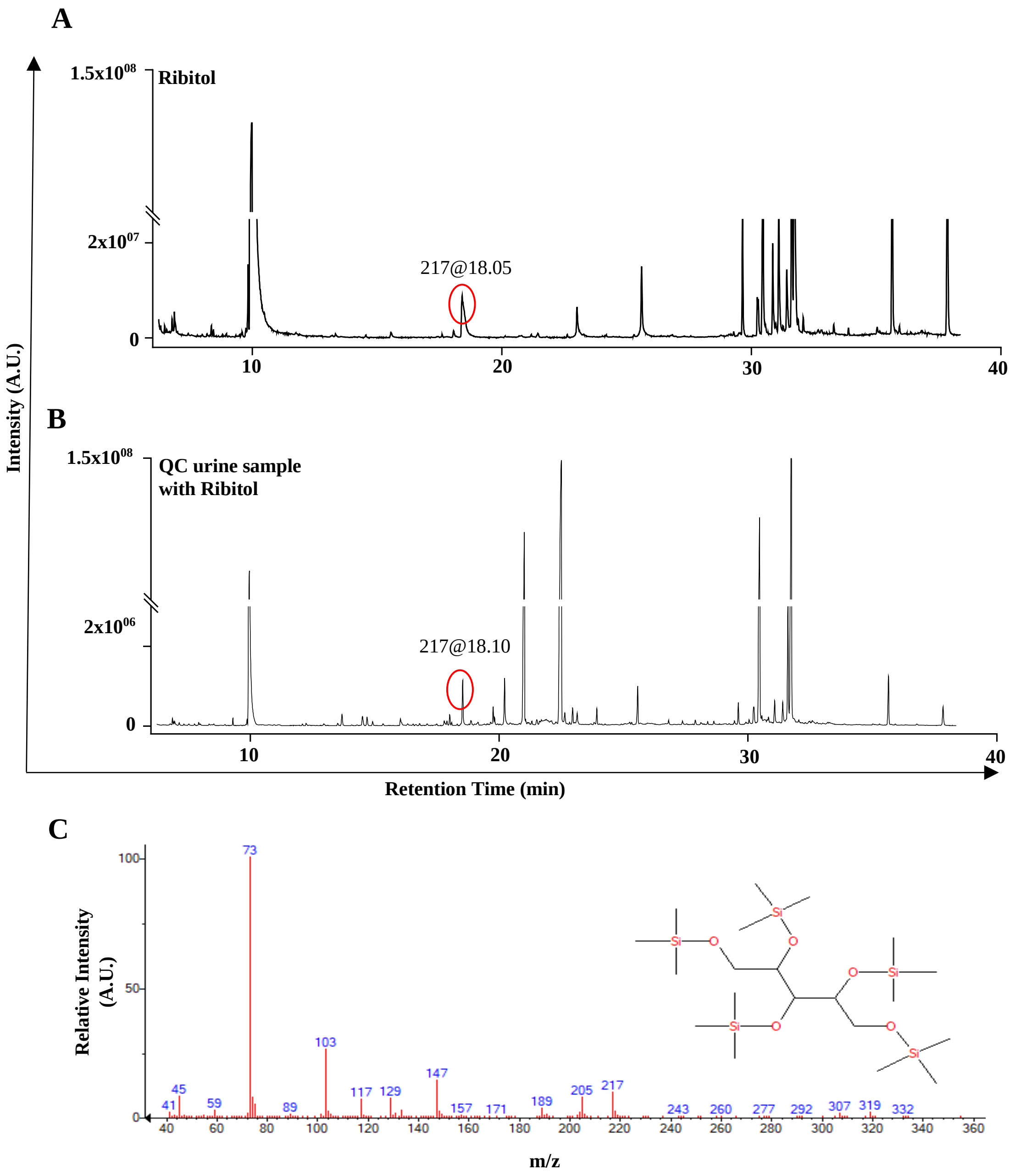

**Figure S1.** Identification of Ribitol (internal standard, 100 ng) and spiked-in quality control (QC) urine sample. **A.** Total ion chromatogram (TIC) of Ribitol alone. **B**. TIC of Ribitol spiked-in QC urine sample. **C.** Extracted ion chromatogram (XIC) of Ribitol. A.U.: arbitrary units, min: minutes, m/z: mass-to-charge ratio.

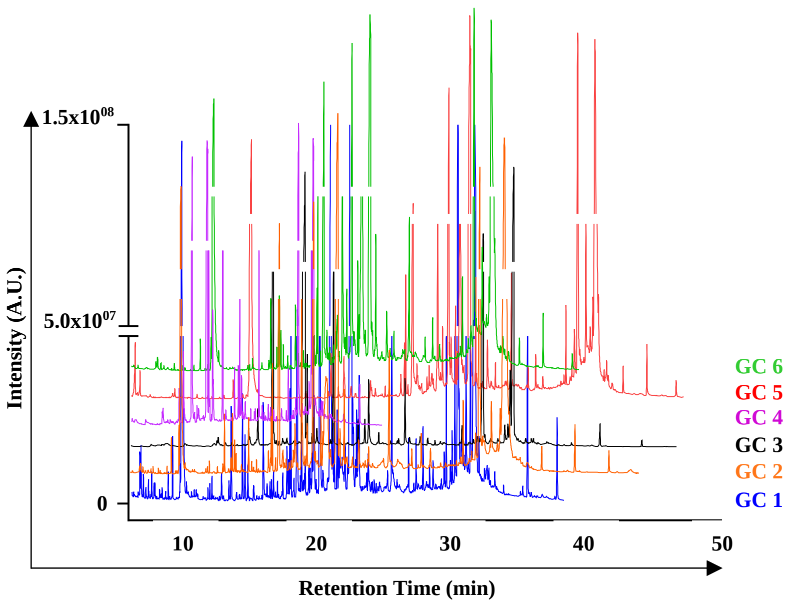

**Figure S2.** Total ion chromatograms (TIC) obtained from different Gas Chromatography and Mass Spectrometry (GC-MS) methods used for global urine metabolite analysis. **GC 1:** 50 °C for 1 min, ramp of 10 °C/min to 150 °C, hold for 3 min, ramp of 7 °C/ min to 300 °C, hold for 3 min (run time 38.43 min); **GC 2:** 50 °C for 1 min, ramp of 10 °C/min to 150 °C, ramp of 5 °C/ min to 300 °C, hold for 3 min (run time 44 min; **GC 3:** 120 °C for 0 min, ramp of 4 °C/min to 300 °C, hold for 2 min (run time 47 min); **GC 4:** 120 °C for 0 min, ramp of 8 °C/min to 300 °C, hold for 2.5 min (run time 25 min); **GC 5:** 50 °C for 1 min, ramp of 5 °C/min to 150 °C for 2 min, ramp of 7 °C/ min to 300 °C, hold for 3 min (run time 47.43 min); **GC 6:** 50 °C for 1 min, ramp of 7 °C/min to 300 °C, hold for 3 min (run time 39.71 min). A.U.: arbitrary units, min: minutes.

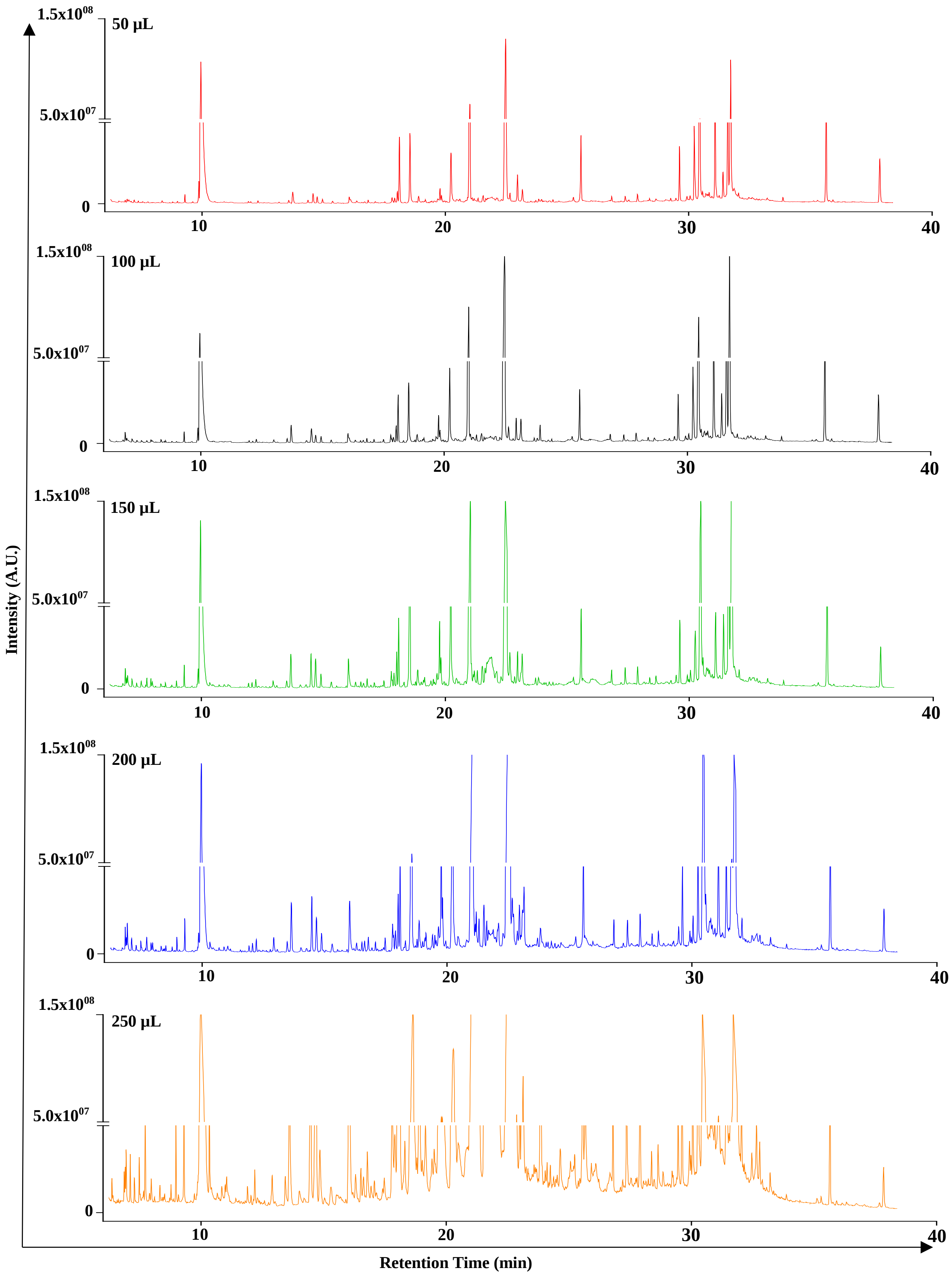

**Figure S3.** Total ion chromatogram showing the peak distribution of different quality control urine sample volumes used for metabolite extraction, derivatization and Gas Chromatography-Mass Spectrometry data acquisition. A.U.: arbitrary units, min: minutes.

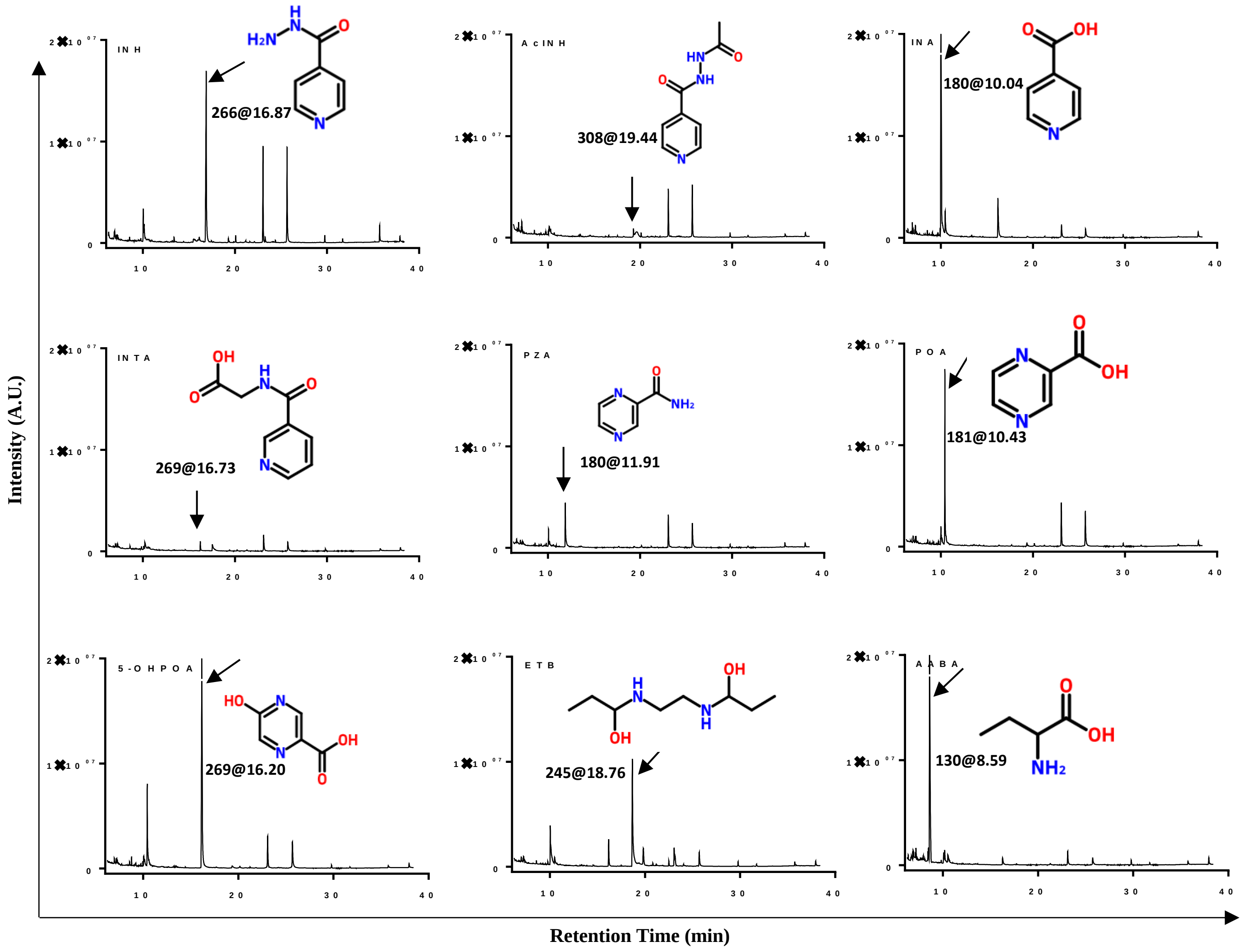

**Figure S4.** Total ion chromatogram and retention time of the derivatized commercial standard of parent anti-tuberculosis drugs and their breakdown products as obtained from Gas Chromatography and Mass Spectrometry analysis. A.U.: arbitrary units, min: minutes.

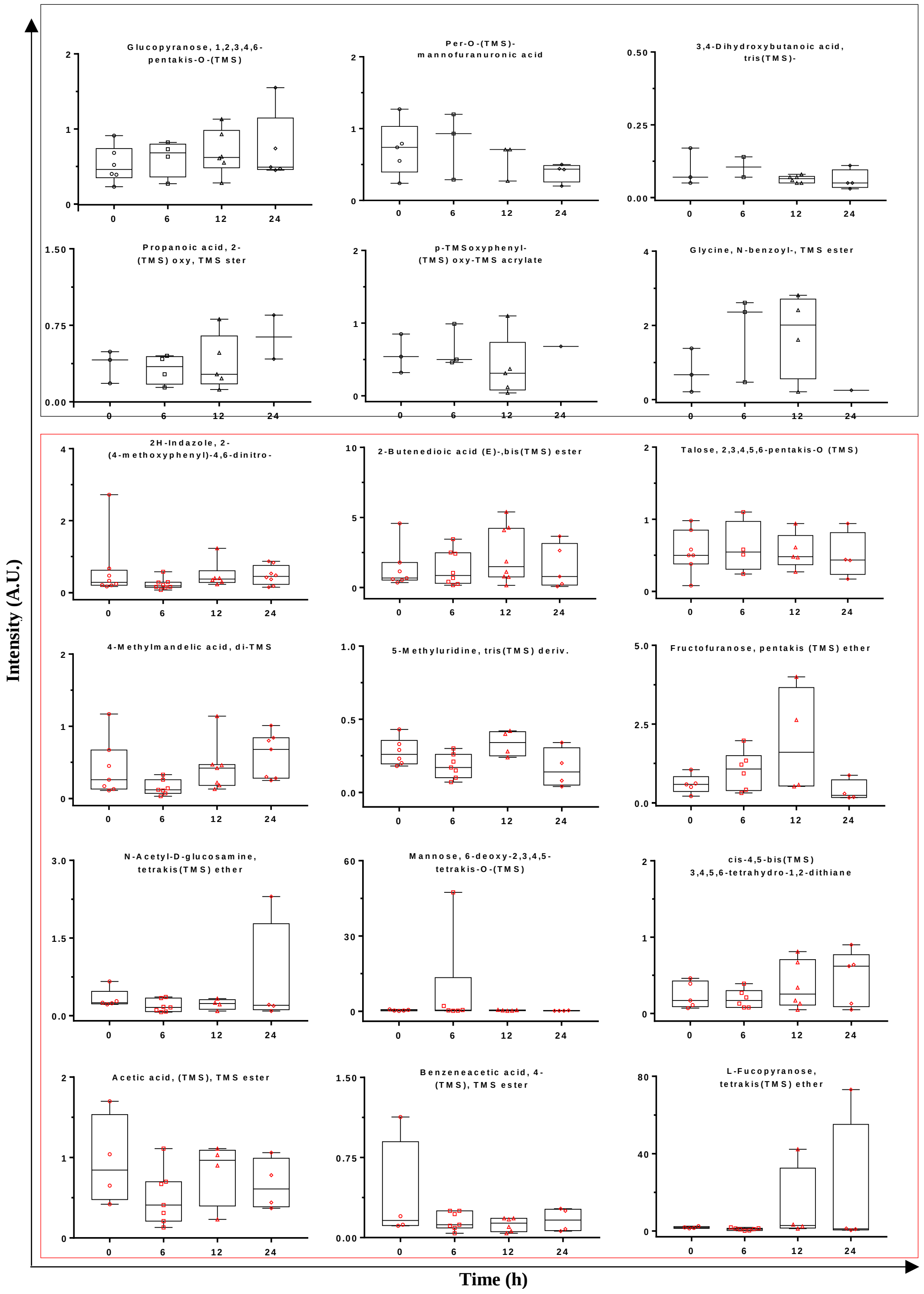

**Figure S5.** Unique urine analytes identified in controls (black) and active tuberculosis (red) subjects showed diurnal variation in their abundance at different times of the day. A.U.: arbitrary units; h: hours.

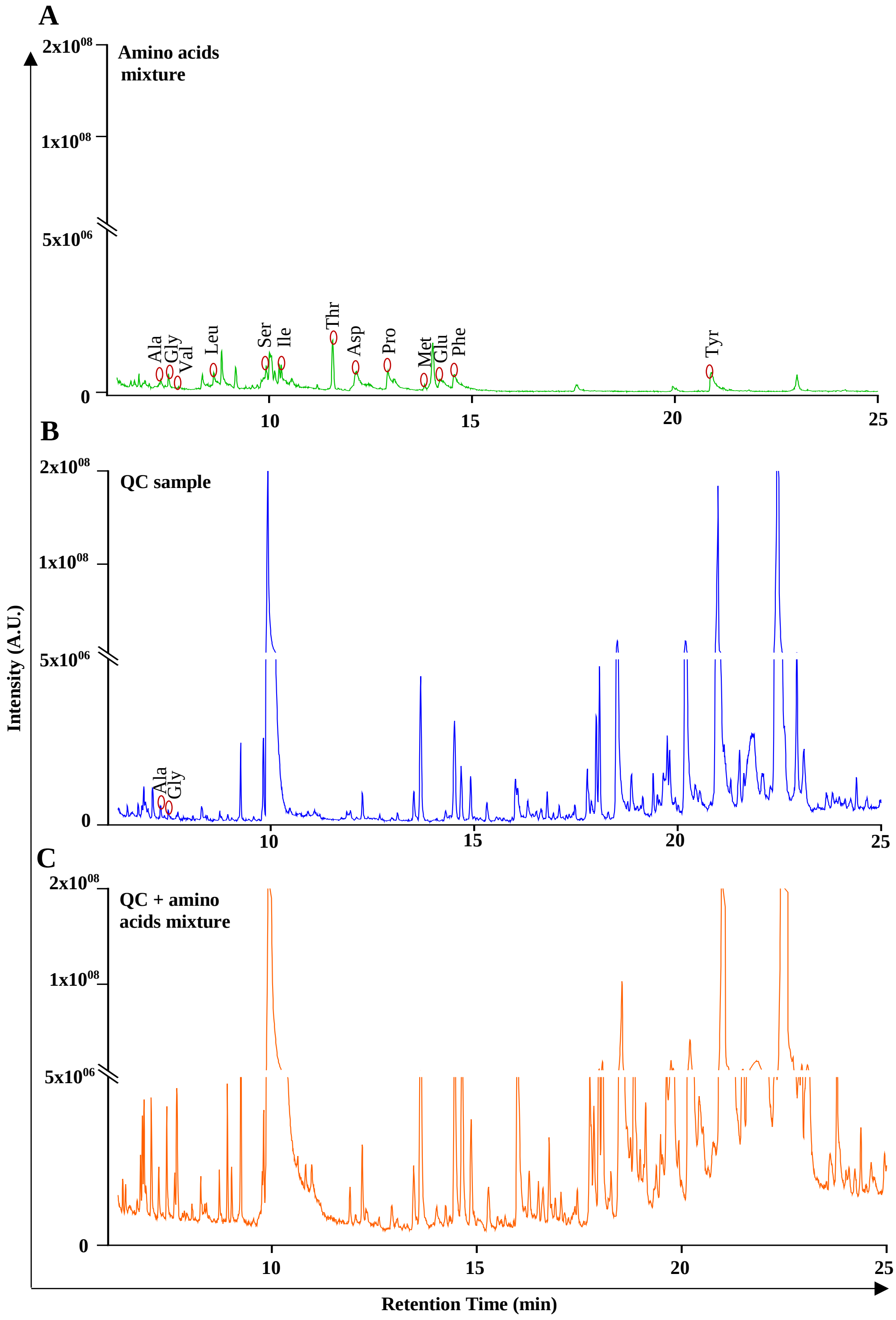

**Figure S6.** Total ion chromatogram (TIC) and retention time of the derivatized commercial standards of amino acids and quality control (QC) urine sample using the optimized GC-MS method; Total run time was 38.43 min and the identified amino acids eluted before 25 min, so for clarity, the X-axis is presented till 25 min here. **A.** TIC of derivatized standard amino acids mixture; Derivatized identified amino acids (e.g., Ala-TMS, Gly-TMS, etc.) are represented with their names only. **B.** TIC of identified amino acids in QC urine sample. **C.** TIC of amino acids spiked-in QC urine sample. A.U.: arbitrary units, min: minutes.

**Table S1:** Epidemiological details of study subjects. (ATB: active tuberculosis patients)

| Parameters | Study Groups | | | |
|---|---|---|---|---|
| | **Total** | **Healthy** | **ATB** | |
| **Demographic parameters** | | | | |
| Sex (Male/Female) | 10/5 | 4/3 | 6/2 | |
| Mean Age (range) in years | 37 (24-70) | 29.3 (24-35) | 43.7 (25-70) | $p = 0.0037$ |
| Body Mass Index ($kg/m^2$) | 22.5 ±3.3 | 24.0±2.8 | 21.2±3.3 | $p = 0.0072$ |
| Smoker (%) | 53 | 29 | 75 | |
| Alcohol user (%) | 40 | 29 | 50 | |

**Table S2:** Tentatively identified human urine analytes in healthy control and active tuberculosis patients receiving treatment.

| S. No. | Analyte No. | **Analyte Name** | **Class** |
|---|---|---|---|
| 1 | M66 | Propanamide, 2-(1,3-dihydro-1,3-dioxo-2H-isoindol-2-yl)-N-[2-(4-propylphenoxy) ethyl]- | Amide |
| 2 | M59 | N-Acetyl-D-glucosamine, tetrakis (TMS) ether | |
| 3 | M22 | Alanine, N-(TMS)-, TMS ester | Amino acid |
| 4 | M43 | Glycine, N-(TMS)-, trimethylsilyl ester | |
| 5 | M44 | Glycine, N-[4-[(TMS-oxy) benzoyl]-, TMS ester | |
| 6 | M45 | Glycine, N-benzoyl-, TMS ester | |
| 7 | M51 | L-Leucine, N-(TMS)-, TMS ester | |
| 8 | M52 | L-Proline, 5-oxo-1-(TMS)-, TMS ester | |
| 9 | M58 | N, O-Bis-(TMS)valine | |
| 10 | M82 | N-(TMS) pyrazine-2-carboxamide | Anti-TB drugs derivative |
| 11 | M81 | 3-Aminoisobutyric acid, 2TMS | |
| 12 | M83 | 3-Pyridinecarboxylic acid, TMS ester | |
| 13 | M84 | Pyrazine-2-carboxylic acid, TMS ester | |
| 14 | M85 | Pyrazine-3,6-dimethyl-2,5, -bis(trimethylsiloxy) | |
| 15 | M3 | 1-Cyano-6-trifluoromethylphenanthrene | Aromatic hydrocarbon |
| 16 | M9 | 2,3-Dihydroxybutanoic acid, tris (TMS)- | Fatty acid |
| 17 | M15 | 3,4-Dihydroxybutanoic acid, tris (TMS)- | |
| 18 | M46 | Hexadecanoic acid, TMS ester | |
| 19 | M60 | Octadecanoic acid, 2,3-bis [TMS-oxy] propyl ester | |
| 20 | M61 | Octadecanoic acid, TMS ester | |
| 21 | M23 | Androst-16-ene-17-carbonitrile | Hormone |
| 22 | M7 | 1-Trimethylsiloxy-2-TMS aminoethane | Hydrocarbon |
| 23 | M8 | 2,2-Bis[4'-cyanooxyphenyl] propane | |
| 24 | M5 | 1-Methyl-N, N-bis (TMS)-4-[(TMS)oxy]-1H-imidazol-2-amine | Imidazole derivative |
| 25 | M4 | 1H-Indole-2,3-dione, 1-(TMS)-, 3-(O-ethyloxime) | Indole quinone |
| 26 | M64 | Phosphoric acid, bis (TMS) 2,3-bis [TMS-oxy] propyl ester | |

| | | | |
|---|---|---|---|
| 27 | M28 | Bis(tert-butyldimethylsilyl) carbonate | Inorganic acid |
| 28 | M13 | 2-Methylacetoacetic acid, di (TMS) deriv. | Keto acid |
| 29 | M33 | Fructofuranose, pentakis (TMS) ether | Monosaccharide |
| 30 | M35 | Galactopyranoside, methyl 2,3,6-tris-O-(TMS)-, acetate | |
| 31 | M36 | Galactopyranoside, methyl 2,3-bis-O-(TMS)-, cyclic phenylboronate | |
| 32 | M38 | Galactose, 2-(acetylamino)-2-deoxy-3,4,5,6-tetrakis-O-(TMS) | |
| 33 | M39 | Glucopyranose, 1,2,3,4,6-pentakis-O-(TMS) | |
| 34 | M47 | Hexopyranose, 1,2,3,4,6-pentakis-O-(TMS) | |
| 35 | M49 | L-Fucopyranose, tetrakis (TMS) ether | |
| 36 | M50 | L-Fucose, tetrakis (TMS) ether | |
| 37 | M55 | Lyxopyranose, tetrakis (TMS) ether | |
| 38 | M56 | Mannose, 6-deoxy-2,3,4,5-tetrakis-O-(TMS)-, L- | |
| 39 | M68 | Psicose, pentakis (TMS) ether | |
| 40 | M76 | Talose, 2,3,4,5,6-pentakis-O-(TMS)- | |
| 41 | M80 | Xylulose tetrakis (TMS)- | |
| 42 | M19 | 5-Methyluridine, tris (TMS) deriv. | Nucleoside base |
| 43 | M1 | 1,2,3-Propanetricarboxylic acid, 2-[(TMS) oxy]-, tris (TMS) ester | Organic acid |
| 44 | M6 | 1-Propene-1,2,3-tricarboxylic acid, tris (TMS) ester, (E)- | |
| 45 | M10 | 2-Butenedioic acid (E)-, bis (TMS) ester | |
| 46 | M14 | 2-Propenoic acid, 2-[(TMS)oxy]-, TMS ester | |
| 47 | M16 | 3-Hydroxy-3-(4'-hydroxy-3'-methoxyphenyl) propionic acid, tri-TMS | |
| 48 | M17 | 4-Methylmandelic acid, di-TMS | |
| 49 | M20 | Acetamide, N-(2,5-dichlorophenyl)-2-oxo-2-[N2-(1-phenylpropyliden) hydrazino]- | |
| 50 | M21 | Acetic acid, [(TMS)oxy]-, TMS ester | |
| 51 | M24 | Arabino-Hexonic acid, 3-deoxy-2,4,5,6-tetrakis-O-(TMS)-, TMS ester | |
| 52 | M25 | Benzeneacetic acid, 4-[(TMS)oxy]-, TMS ester | |
| 53 | M26 | Benzenepropanoic acid,4-bis[(TMS)oxy]-, TMS ester | |
| 54 | M29 | Butanedioic acid, methylene-, bis (TMS) ester | |

| | | | |
|---|---|---|---|
| 55 | M30 | Butanoic acid, 2,4-bis[(TMS)oxy]-, TMS ester | |
| 56 | M57 | N, N-Diethyl (TMS)carbamate | |
| 57 | M67 | Propanoic acid, 2-[TMS-oxy]-, TMS ester | |
| 58 | M69 | p-Trimethylsilyloxyphenyl-(TMS-oxy) TMS acrylate | |
| 59 | M75 | Succinic acid, 2,3-bis(trimethylsiloxy)-, bis (TMS) ester | |
| 60 | M63 | Phosphine oxide, bis (pentamethyl phenyl)- | Organophosphine |
| 61 | M78 | Trp-Arg | Peptide |
| 62 | M41 | Glycerol, tris (TMS) ether | Polyol |
| 63 | M65 | Pinitol, pentakis (TMS) ether | |
| 64 | M12 | 2-Keto-l-gluconic acid, penta (O-TMS)- | Sugar acid |
| 65 | M40 | Glucuronic acid, 2,3,4,5-tetrakis-O-(TMS)-, TMS ester | |
| 66 | M54 | L-Threonic acid, tris (TMS) ether, TMS ester | |
| 67 | M73 | Ribonic acid, 2,3,4,5-tetrakis-O-(TMS)-, TMS ester | |
| 68 | M74 | Ribonic acid, 2-desoxy-tetrakis-O-(TMS)- | |
| 69 | M71 | Ribitol, 1,2,3,4,5-pentakis-O-(TMS)- | Sugar acid/ Internal Standard |
| 70 | M2 | 1,5-Anhydro-D-sorbitol, tetrakis (TMS) ether | Sugar alcohol |
| 71 | M53 | L-Threitol, tetrakis (TMS) ether | |
| 72 | M79 | Xylitol, 1,2,3,4,5-pentakis-O-(TMS)- | |
| 73 | M27 | Benzo [d, E] isoindolo[2,1-a] quinazolin-12-one, 11-amino- | Not grouped |
| 74 | M42 | Glycero-L-manno-Heptonic acid, 2,3,5,6,7-pentakis-O-(TMS) lactone | |
| 75 | M18 | 5-[Cyano-(3,4-dimethyl-5-oxo-1,5-dihydro-pyrrol-2-ylidene)-methyl]-2,3,3-trimethyl-3,4-dihydro-2H-pyrrole-2-carbonitrile | |
| 76 | M31 | cis-4,5-bis (TMS-oxy)-3,4,5,6-tetrahydro-1,2-dithiane | |
| 77 | M32 | Ethyl 2,3,4,6-tetrakis-O-(TMS)-D-glucopyranoside | |
| 78 | M34 | Galactofuranosiduronic acid, methyl 2,3,5-tris-O-(TMS)-, methyl ester | |
| 79 | M37 | Galactopyranosiduronic acid, methyl 2,3,4-tris-O-(TMS)-, methyl ester | |
| 80 | M48 | Inosose-2, 1,3,4,5,6-pentakis-O-(TMS)-, myo- | |
| 81 | M62 | Per-O-(TMS)-à-d-mannofuranuronic acid | |
| 82 | M70 | Pyrido[3,4-d] pyrimidin-4(3H)-one, 6,8-dimethyl- | |

| | | |
|---|---|---|
| 83 | M72 | Ribo-Hexitol, 3-deoxy-1,2,4,5,6-pentakis-O-(TMS) |
| 84 | M77 | Thiazolo[4,5-f] quinoline, 2,7,9-trimethyl- |
| 85 | M11 | 2H-Indazole, 2-(4-methoxyphenyl)-4,6-dinitro- |
